# Efficacy and Safety of a Novel Dietary Supplement, Gepaktiv (International name Phenomenon), Versus Active Comparators (UDCA and Ademetionine) in Patients with Metabolic-Associated Fatty Liver Disease: A Preliminary Comparative Analysis

**DOI:** 10.1101/2025.08.04.25332290

**Authors:** Evgeniy V. Chesnokov

## Abstract

**Background:** Metabolic-associated fatty liver disease (MAFLD) is a prevalent chronic liver condition with limited approved pharmacological treatments [1]. Ursodeoxycholic acid (UDCA) and ademetionine show variable efficacy, primarily on liver enzymes. This study presents a preliminary analysis comparing the efficacy of a novel dietary supplement, Gepaktiv, against these comparators in MAFLD patients.

**Methods:** In this open-label, randomized controlled trial (clinicaltrials.gov NCT07068191 and ITMCTR 2025001469), 19 patients with MAFLD, confirmed by hepatomegaly (liver size ≥3 cm above normal by ultrasound), elevated alanine aminotransferase (ALT, 90–150 U/L), and FibroScan results (steatosis ≥260 dB/m, fibrosis ≥11 kPa), were allocated to Gepaktiv (n=6, 1500 mg/day), UDCA (n=7, 10 mg/kg/day), or Ademetionine (n=6, orally 400 mg 2 times a day) for 15 days. Patients with significant alcohol consumption (>20 g/day for women, >30 g/day for men) were excluded. Primary outcomes were median changes from baseline to day 15 in ALT, aspartate aminotransferase (AST), liver size (craniocaudal diameter, cm, via ultrasound), steatosis (controlled attenuation parameter, CAP, dB/m), and fibrosis (transient elastography, kPa).

**Results:** The Gepaktiv group showed median [IQR] reductions of ALT -48.9 [-54.0 to -35.0] U/L, AST - 62.8 [-66.0 to -44.0] U/L, liver size -1.9 [-2.0 to -1.2] cm, and steatosis -32.5 [-45.0 to -30.0] dB/m. These reductions were significantly greater compared to both UDCA and Ademetionine groups (p < 0.01 for ALT, AST, and liver size; p < 0.05 for steatosis). Fibrosis reduction was minimal and not statistically significant between groups.

**Conclusion:** The Gepaktiv group was associated with greater improvements in biochemical and imaging markers of MAFLD compared to UDCA and Ademetionine in this preliminary analysis. These findings warrant further investigation in larger, long-term trials.

*Note*: These preliminary results have not been peer-reviewed and should not guide clinical practice.

## Introduction

Metabolic-associated fatty liver disease (MAFLD), affecting approximately 25% of the global population, encompasses a spectrum from simple steatosis to metabolic-associated steatohepatitis (MASH), with risks of progression to cirrhosis and hepatocellular carcinoma [1]. Unlike the previous non-alcoholic fatty liver disease (NAFLD) framework, MAFLD emphasizes metabolic dysfunction, including overweight/obesity, type 2 diabetes, or metabolic syndrome, as diagnostic criteria [1]. Lifestyle interventions, including diet and exercise, remain the cornerstone of management, but pharmacological options are limited. Ursodeoxycholic acid (UDCA) and ademetionine, commonly used as hepatoprotective agents, show modest effects on liver enzymes and minimal impact on steatosis or fibrosis [2,3]. Gepaktiv, a novel dietary supplement containing milk thistle extract [4, 8], pumpkin, agrimony, nettle, chamomile, and other components, has shown preclinical promise in reducing oxidative stress and hepatic fat accumulation. This preliminary analysis from a randomized controlled trial (GEP-2025-01) evaluates the short-term efficacy and safety of Gepaktiv compared to UDCA and Ademetionine in patients with MAFLD. The rationale for this clinical trial is based on significant preclinical findings. In a CCl4-induced cirrhosis model in rats, Phenomenon (Gepaktiv) demonstrated a potent therapeutic effect, significantly improving liver functional parameters (ALT, AST), normalizing indicators of oxidative stress, and reducing liver fibrosis

## Methods

### Study Design and Participants

This open-label, randomized controlled trial with blinded outcome assessment was conducted at Tyumen State Medical University, Russia. Patients aged 18–65 with MAFLD, confirmed by hepatomegaly (liver size ≥3 cm above normal by ultrasound), elevated ALT (90–150 U/L), and FibroScan results indicating steatosis (CAP ≥260 dB/m) and fibrosis (≥11 kPa), were enrolled. Exclusion criteria included cirrhosis, malignancy, pregnancy, allergies, gallstones, liver atrophy, liver cysts and liver nodules. All patients adhered to a standardized diet excluding dairy, eggs, pork, lamb, mayonnaise, alcohol, and narcotics, monitored via patient diaries.

Randomization was performed by the study coordinator using a 1:1:1 block method and a random number generator to Gepaktiv, UDCA, or Ademetionine. Outcome assessors (imaging specialists (including radiologists and trained sonographers), trained FibroScan operators, and laboratory staff) were blinded to group allocation.

### Outcomes

The **primary outcomes** were the median changes from baseline to day 15 in the following parameters:

- Serum Alanine Aminotransferase (ALT) level (U/L).
- Serum Aspartate Aminotransferase (AST) level (U/L).

The **key secondary outcomes** included:

- The proportion of patients achieving a therapeutic response, defined as a reduction in ALT level of ≥30% from baseline, a validated surrogate marker for histological improvement [7].
- The median change from baseline in liver steatosis, as measured by the Controlled Attenuation Parameter (CAP, dB/m).
- The median change from baseline in liver size, as measured by the craniocaudal diameter (cm) via ultrasound.
- The median change from baseline in liver stiffness, as a measure of fibrosis (kPa).

Additional exploratory parameters (e.g., gamma-glutamyl transferase, bilirubin, CLDQ quality-of-life score) were collected but are not reported in this interim analysis.

This manuscript has not been peer-reviewed or submitted for publication elsewhere.

### Interventions

#### Participants received

Gepaktiv: (codename: Phenomenon), manufactured by Phenomenon Pharma LLC, Russia (Lot No. 03250327). Each 250 mg capsule contains a proprietary formulation consisting of microcrystalline cellulose (anti-caking agent), gelatin capsule shell (with titanium dioxide, quinoline yellow dyes), Silybum marianum (milk thistle) fruit extract, sublimated pumpkin, Agrimonia eupatoria (agrimony) extract, Urtica dioica (nettle) leaf extract, ferrous sulfate, Matricaria chamomilla (chamomile) flower extract, methylsulfonylmethane (MSM), Aralia mandshurica extract, and Schisandra chinensis extract. ^**^According to the information provided by the manufacturer (Phenomenon Pharma LLC)^**^, the components are processed using an exclusive proprietary technology designed to enhance the bioavailability and synergistic activity of the active substances. Therefore, the therapeutic effects described in this study are hypothesized to be specific to this particular formulation and manufacturing process.

UDCA: Patients randomized to the UDCA group were instructed to purchase and self-administer ursodeoxycholic acid, available over-the-counter or by prescription in local pharmacies, at a standard therapeutic dose of 10 mg/kg/day. As the medication was sourced by the participants themselves, the specific brand and manufacturer were not standardized or controlled by the study investigators.

Ademetionine: Similarly, patients in the Ademetionine group were instructed to purchase and self-administer ademetionine at a dose of orally 400 mg 2 times a day. The specific brand and manufacturer of ademetionine were not standardized as the product was procured by the participants.

Treatment duration was 15 days, with assessments at baseline (day 1), day 7 (data not reported), and day 15.

## Statistical Analysis

Due to the small sample sizes and the non-normal distribution of the data, non-parametric statistical methods were employed. Data are presented as Median [Interquartile Range (IQR)]. Between-group comparisons were performed using the Mann-Whitney U test. A p-value of less than 0.05 was considered statistically significant. All analyses were performed using software GraphPad Prism 9.

## Results

This interim analysis includes data from 19 patients who successfully completed the 15-day study protocol: Gepaktiv (n=6), UDCA (n=7), and Ademetionine (n=6). Adherence to the study intervention and diet was high across all groups.

The Gepaktiv group demonstrated clinically and statistically significant improvements across most endpoints compared to the active control groups. The results are presented as medians and interquartile ranges to accurately reflect the data distribution. A summary of the changes from baseline to day 15 is presented in Table 1.

**Table 1.** Median Change [IQR] in Liver Parameters After 15 Days of Treatment.

| Parameter | Gepaktiv (n=6) | UDCA (n=7) | Ademetionine (n=6) |
| --- | --- | --- | --- |
| ALT Reduction (U/L) | -48.9 [-54.0 to -35.0] | -14.0 [-15.0 to -6.0] | -17.0 [-24.0 to -14.0] |
| AST Reduction (U/L) | -62.8 [-66.0 to -44.0] | -17.7 [-30.0 to -10.0] | -22.5 [-27.0 to -17.0] |
| Reduction in Liver Span (cm) | -1.9 [-2.0 to -1.2] | 0.0 [0.0 to 0.0] | -0.05 [-0.2 to 0.0] |
| Steatosis (CAP) Reduction (dB/m) | -32.5 [-45.0 to -30.0] | -3.0 [-6.0 to -2.0] | -2.5 [-7.0 to -2.0] |
| Fibrosis Reduction (kPa) | -0.2 [-0.3 to -0.1] | -0.1 [-0.1 to 0.0] | -0.1 [-0.2 to 0.0] |

Statistical comparisons using the Mann-Whitney U test confirmed that the reductions observed in the Gepaktiv group were significantly greater than those in both the UDCA and Ademetionine groups for the key parameters of ALT, AST, liver span, and steatosis (CAP). The results of the between-group comparisons are detailed in **Table 2**.

**Table 2.**
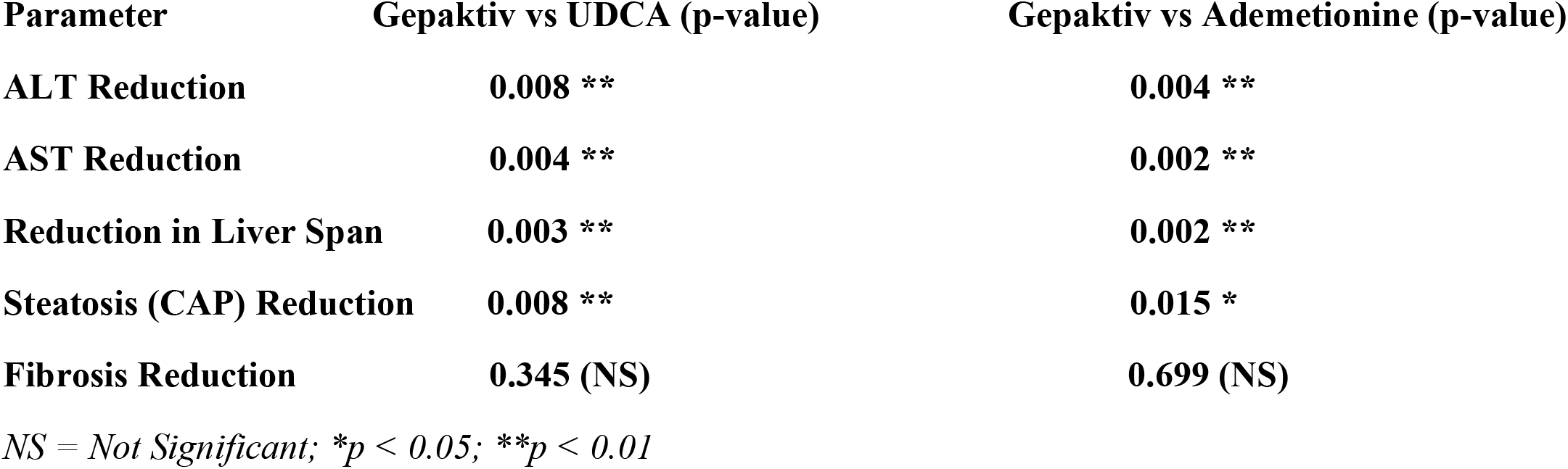
Statistical Comparison of Treatment Effects (Mann-Whitney U test)

| Parameter | Gepaktiv vs UDCA (p-value) | Gepaktiv vs Ademetonine (p-value) |
| --- | --- | --- |
| ALT Reduction | 0.008 ** | 0.004 ** |
| AST Reduction | 0.004 ** | 0.002 ** |
| Reduction in Liver Span | 0.003 ** | 0.002 ** |
| Steatosis (CAP) Reduction | 0.008 ** | 0.015 * |
| Fibrosis Reduction | 0.345 (NS) | 0.699 (NS) |

NS = Not Significant; \**p* < 0.05; \*\**p* < 0.01

The magnitude of these differences is visualized in Figure 1. Data visualization suggests a distinct separation in treatment effects for the key clinical outcomes. This was particularly evident in the reduction of liver enzymes (ALT, AST), the decrease in liver span, and the substantial improvement in steatosis, all strongly in favor of Gepaktiv. Consistent with the short 15-day duration of this initial analysis, the median changes in liver stiffness (fibrosis) were minimal and did not differ significantly between the groups. The assessment of this parameter over a longer duration is planned for the 30- and 60-day follow-up points.

**Fig. 1.**
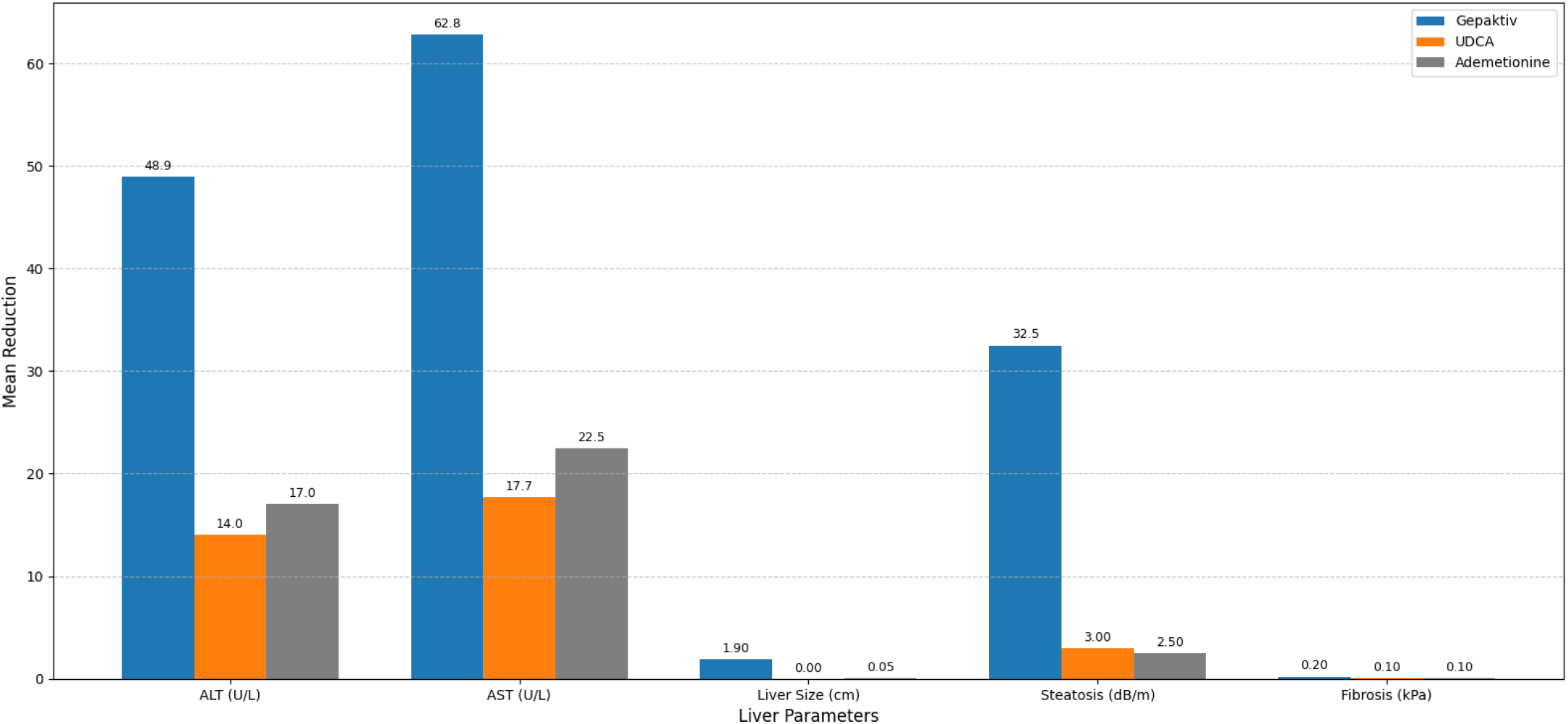

## Discussion

The Gepaktiv group was associated with greater reductions ALT (-48.9U/L), AST (-62.8 U/L), liver size (- 1.9 cm), steatosis (-32.5 dB/m), and fibrosis (-0.2 kPa) compared to UDCA and Ademetionine over 15 days [2, 5, 8]. These preliminary observations are consistent with preclinical data suggesting Gepaktiv’s antioxidant, anti-inflammatory, and lipid-lowering properties, potentially driven by milk thistle and agrimony [8]. The reductions in ALT and AST exceeded the protocol’s surrogate marker threshold (≥30% reduction), indicating improved liver function [7]. Preliminary evidence suggests a significant reduction in steatosis, indicating a potential effect on hepatic fat metabolism, a key feature of MAFLD [1,4].

The modest effects of UDCA and Ademetionine are consistent with prior studies showing limited impact on steatosis and fibrosis [2,5,6]. The small sample size (n=19) and short duration (15 days) limit generalizability, and potential confounders (e.g., baseline disease severity, CAP measurement variability) were not assessed due to the interim nature of this analysis [9]. The lack of significant change in liver stiffness at day 15 is an expected finding, as fibrosis is a structural parameter that requires a longer period to demonstrate measurable improvement. However, given the potent effects observed on steatosis and liver enzymes in this study, and supported by anti-fibrotic preclinical data, we hypothesize that a longer treatment duration is necessary to reveal a significant impact on this outcome. This hypothesis will be directly tested at the upcoming 30- and 60-day assessments. The full study (n=90) with extended follow-up (30–60 days) will provide further insights. The standardized diet, monitored via diaries, may have contributed to improvements across groups, though Gepaktiv’s superior outcomes suggest a treatment-specific effect [10].

The composition of Gepaktiv, consisting of natural ingredients, does not represent a fundamentally new set of components compared to other hepatoprotective agents. However, the observed efficacy is likely due to an exclusive proprietary manufacturing technology that optimizes the bioavailability and therapeutic activity of the active substances. This technology, protected as a commercial secret or patent, is presumed to enhance the action of the natural components, distinguishing Gepaktiv from its analogs.

## Ethics

The study was approved by the Tyumen State Medical University Ethics Committee (Protocol № 131, 30 June 2025) and conducted in accordance with the Declaration of Helsinki, ICH Good Clinical Practice, and Russian regulations (FZ-323, FZ-152, GOST R 52379-2005). Informed consent was obtained from all participants.

## Safety

Treatment was well-tolerated across all three groups. No serious adverse events or adverse events leading to discontinuation of the study intervention were reported during the 15-day observation period.

## Conclusion

This preliminary analysis suggests Gepaktiv offers greater improvements in biochemical and imaging markers of MAFLD compared to UDCA and Ademetionine. These findings support further investigation in the ongoing trial (clinicaltrials.gov NCT07068191 and ITMCTR 2025001469) to confirm efficacy and safety in a larger cohort.

## Data Availability

Data supporting the findings of this study are available from the corresponding author, Evgeniy V. Chesnokov, upon reasonable request.

https://clinicaltrials.gov/study/NCT07068191

## Acknowledgments

We thank the study participants, clinical staff, and coordinator Irina M. Nagibina, Tyumen State Medical University, Tyumen, Russia.

## Conflicts of Interest

This study was funded in full by Phenomenon Pharma LLC, the manufacturer of the investigational product Gepaktiv. Tyumen State Medical University received funding from Phenomenon Pharma LLC to conduct the clinical trial. Evgeniy V. Chesnokov, MD, Prof., served as the Principal Investigator for this study.

## Funding

This work was supported by a research grant from Phenomenon Pharma LLC. The funder (Phenomenon Pharma LLC) provided the investigational product (Gepaktiv) and financial support for the conduct of the research but had no role in the design of the study, the collection, analysis, or interpretation of data, the writing of the manuscript, or in the decision to submit the manuscript for publication.

## Data Availability

Data are available from the corresponding author upon reasonable request.

## Notes

### Competing Interest Statement

The author declare a competing interest. This study was funded by Phenomenon Pharma LLC, the manufacturer of the investigated product Gepaktiv (Phenomenon). The funding entity had no influence on the study design, data collection, analysis, interpretation, or manuscript preparation. The author confirms his independence in conducting the research and presenting the results.

### Clinical Trial

NCT07068191, ITMCTR2025001469

### Clinical Protocols

https://clinicaltrials.gov/study/NCT07068191

### Author Declarations

The Tyumen State Medical University Ethics Committee gave ethical approval for this work (Protocol № 131, 30 June 2025).

